# From CCTA to Surgical Strategy: An Integrated AI Framework for Patient-Specific Coronary artery bypass grafting Planning

**DOI:** 10.64898/2026.05.28.26354400

**Authors:** Mostafa Rezaeitaleshmahalleh, Shahab Masoumi, Emeline Debalme, Thor Sundt, Sary F Aranki, Borami Shin, Farhad R. Nezami

## Abstract

**Background:** Coronary artery bypass grafting (CABG) remains the standard of care for complex multivessel and left main coronary artery disease. However, current preoperative planning remains largely subjective, relying on qualitative interpretation of coronary CT angiography (CCTA), operator-dependent stenosis grading, and fragmented multi-software workflows. Invasive fractional flow reserve (FFR), the reference standard for physiologic lesion assessment, is infrequently acquired preoperatively, leaving distal anastomosis planning without an objective hemodynamic basis.

**Methods:** We developed a fully automated, AI-powered platform that converts routine CCTA into a patient-specific CABG planning workflow through five integrated modules: nnU-Net based segmentation of coronary lumen and calcification; quantitative morphological and topological characterization generating more than thirty descriptors; automated stenosis detection using a local reference-radius formulation; a nine-point composite scoring framework for distal anastomosis site selection incorporating luminal caliber, landing-zone length, calcification burden, distal perfusion reserve, and bifurcation proximity; and interactive virtual graft construction coupled to a distributed reduced-order solver for pre- and post-bypass FFR estimation.

**Results:** Lumen segmentation achieved a mean Dice similarity coefficient of 0.96 ± 0.01, whereas calcium segmentation achieved 0.73 &[plusmn] 0.15 on the held-out cohort. Platform-derived FFR demonstrated strong agreement with invasively measured FFR (r=0.96, R^2^=0.9, mean absolute relative difference 1.73 &[plusmn]1.42%) across the evaluated lesions, supporting the physiologic validity of the reduced-order hemodynamic solver. End-to-end analysis from raw CCTA to hemodynamic assessment and virtual graft planning was completed in approximately seven minutes per case on a standard workstation, representing a substantial reduction in processing time compared with conventional multi-tool and CFD-based workflows.

**Conclusions:** The proposed platform demonstrates the feasibility of rapid, reproducible, and physiology-informed CABG planning using routine CCTA. By integrating anatomical characterization, automated target-site analysis, virtual graft construction, and reduced-order hemodynamic assessment into a single workflow, the framework provides objective, quantitative surgical decision support compatible with routine clinical workflows.

## 1. Introduction

Coronary artery bypass grafting (CABG) remains the standard of care for patients with complex coronary artery disease, including multivessel disease, left main stenosis, and concomitant diabetes mellitus, with landmark randomized trials confirming its durable superiority over percutaneous coronary intervention in reducing long-term mortality, myocardial infarction, and repeat revascularization [1, 2]. Isolated CABG continues to be the most commonly performed cardiac surgical procedure in the United States, with more than 160,000 operations captured annually in the Society of Thoracic Surgeons Adult Cardiac Surgery Database [3]. As the complexity of referred patients increases, the demands placed on pre-operative surgical planning have grown correspondingly.

Yet despite this clinical importance, preoperative planning remains largely subjective and experience-dependent. Current workflows rely on visual interpretation of coronary computed tomography angiography (CCTA) or invasive angiography for lesion classification, selection of the anastomosis site, and graft configuration, which is operator-dependent and prone to significant variability between surgeons [4]. Importantly, anatomical stenosis severity does not consistently reflect physiologic significance, limiting the reliability of anatomy-based decision making alone. Invasive fractional flow reserve (FFR), the reference standard for physiologic lesion assessment, is rarely obtained preoperatively due to its cost, procedural risk, and time requirements.

Coronary CTA has the potential to unify anatomical and functional assessments in a single non-invasive acquisition. CT-derived FFR, validated against invasive FFR in the NXT trial and subsequent multicenter studies [5, 6], enables objective hemodynamic lesion grading without catheterization, and the FASTTRACK CABG study demonstrated the feasibility of planning and executing surgical revascularization based entirely on CCTA and FFR-CT [7]. However, translating raw CCTA data into actionable surgical guidance still requires hours of manual segmentation across fragmented multi-tool workflows, rendering routine point-of-care use impractical for most centers.

Recent advances in artificial intelligence have begun to resolve this bottleneck. Deep learning pipelines now achieve coronary lumen segmentation with Dice similarity coefficients exceeding 0.90 and reduced-order hemodynamic models recover clinically relevant pressure and flow distributions at a fraction of the computational cost of full three-dimensional simulation [8, 9]. Nevertheless, an integrated platform unifying these capabilities into a single clinical workflow has not been described.

This study introduces a fully automated, AI-driven platform that converts routine CCTA into a patient-specific CABG planning strategy in approximately seven minutes on a standard workstation. Our aims were to (1) develop and validate an automated segmentation pipeline for simultaneous coronary lumen and calcification delineation; (2) implement quantitative morphological and topological analysis with automated stenosis detection; (3) establish an objective scoring system for optimal distal anastomosis site selection;(4) create an interactive virtual graft generation tool; and (5) integrate a rapid distributed lumped parameter model for pre- and post-bypass hemodynamic assessment.

## 2. Material and methods

### 2.1. Study Population and Data Acquisition

A total of 28 pre-CABG CCTA cases performed at Brigham and Women’s Hospital were retrospectively identified under Institutional Review Board approval (IRB 2018P001524). All study procedures were conducted in accordance with institutional guidelines and the principles outlined in the Declaration of Helsinki. Eligible patients underwent preoperative CCTA studies and had corresponding invasive FFR measurements available for analysis.

### 2.2. Geometric Characterization of coronary vessel and calcium burden

Segmentation of the coronary lumen and calcified plaques was performed using the nnU-Net deep learning framework [10], as detailed in the Supplementary Methods. Full feature definitions for all descriptor families are provided in Supplementary Tables S1 to S5.

Starting from the 3D lumen and calcium segmentation masks, the proposed pipeline first performs topological correction and surface smoothing to generate a watertight triangulated surface mesh. A vessel centerline tree is subsequently extracted through skeletonization and further refined to remove spurious branches and enforce anatomically consistent connectivity. Major coronary territories, including the right coronary artery (RCA), left main coronary artery (LMCA), left anterior descending artery (LAD), and left circumflex artery (LCx), are then automatically identified and labeled based on centerline topology and branching geometry (Fig. 1).

**Figure 1:**
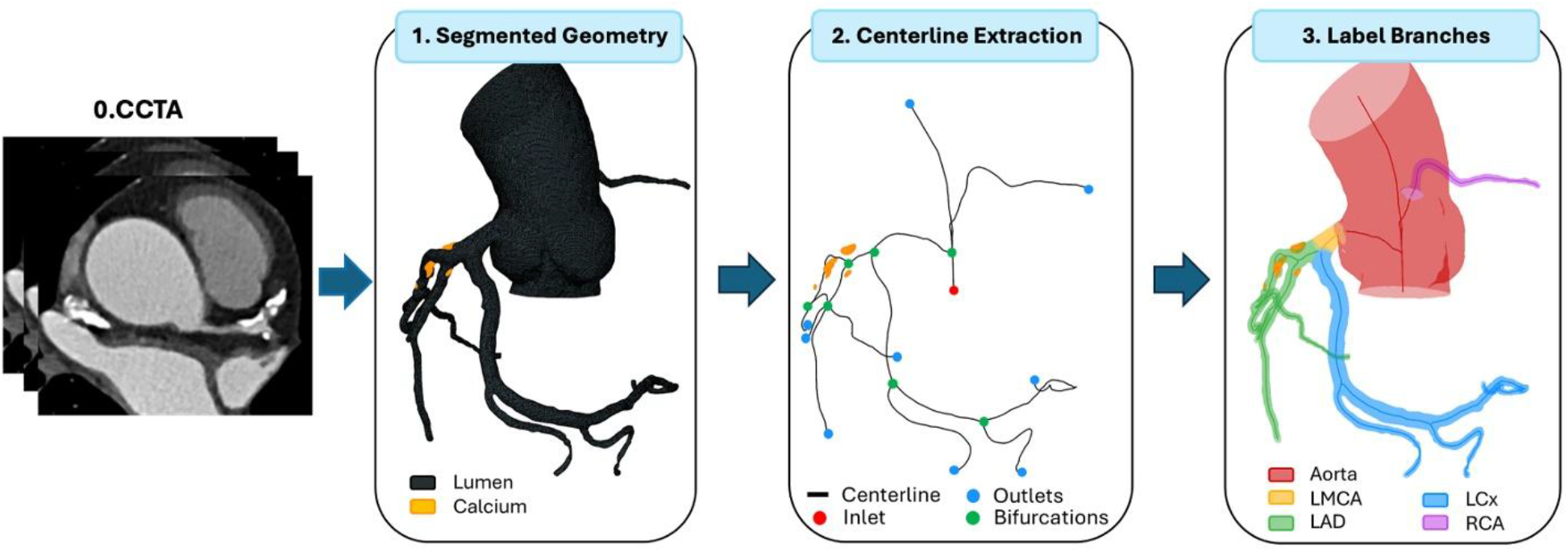
Automated partitioning pipeline: watertight lumen surface mesh, extracted centerline, and anatomical branch labeling for the major coronary territories (RCA, LMCA, LAD, LCx).

#### 2.2.1. Global Calcium Burden

Global coronary calcium burden was quantified using whole-tree metrics derived from the complete calcium segmentation mask. Extracted features included the total number of discrete calcified deposits, cumulative calcification volume, total calcified surface area, and the number of coronary segments containing calcium. Collectively, these descriptors provide a patient-level representation of overall coronary atherosclerotic calcification burden without lesion-specific weighting.

#### 2.2.2. Per-Deposit Calcium Shape

Morphological descriptors were independently computed for each discrete calcified de-posit, including sphericity, elongation, flatness, and mean CT attenuation intensity. To appropriately weight larger and potentially more hemodynamically relevant lesions, features were subsequently aggregated across all deposits using volume-weighted averaging.

#### 2.2.3. Luminal Geometry

Coronary luminal geometry was characterized using centerline-based descriptors computed at the branch level and aggregated across the coronary tree using length-weighted averaging to account for variations in vessel extent. Extracted features included total vessel length, centerline tortuosity, local curvature, torsion, lumen radius profile, and bifurcation angles. Together, these metrics capture the three-dimensional geometric complexity of the coronary vasculature and its potential influence on local flow dynamics.

#### 2.2.4. Calcium Topology

Topological descriptors were designed to characterize the spatial distribution and anatomical localization of coronary calcifications within the vascular tree. Features included spatial sparsity of calcium deposits, proximity to bifurcation points, dominant segment involvement, and localization relative to the myocardial or pericardial vessel wall. These descriptors complement conventional shape-based features by capturing the anatomical context of calcified lesions relative to hemodynamically important vascular landmarks.

#### 2.2.5. Lumen–Calcium Interaction

Lumen–calcium interaction descriptors quantified the spatial relationship between calcified plaque and the adjacent coronary lumen. Extracted features included the minimum distance between calcium and lumen boundaries, lumen contact ratio, and the arc angle subtended by calcified deposits around the local vessel cross-section. Collectively, these metrics provide a mechanistically relevant characterization of plaque-induced luminal encroachment and geometric distortion, complementing conventional stenosis severity assessment.

### 2.3. Automated Distal Anastomosis Site Selection

Selection of an appropriate distal anastomosis target is a critical determinant of long-term graft patency and clinical outcomes following CABG [11]. To support patient-specific surgical planning, an automated framework was developed to identify anatomically favorable distal target regions directly from CCTA-derived coronary anatomy.

The proposed framework uses the processed coronary centerline tree to establish a continuous proximal-to-distal anatomical representation of each vessel branch. Anatomical land-marks, including bifurcation locations and calcified plaque regions, are automatically identified and mapped along the coronary tree. For each detected coronary stenosis, the algorithm searches for potential distal landing zones downstream from the diseased segment within a predefined anatomical window.

Candidate target regions were required to satisfy several clinically motivated criteria, including adequate vessel caliber, sufficient disease-free landing-zone length, minimal adjacent calcification, preserved distal runoff, and appropriate distance from major bifurcations. Each candidate region was evaluated using a composite scoring framework integrating vessel diameter, landing-zone length, calcification burden, distal perfusion reserve, and local bifurcation anatomy (Table 1). Larger-caliber vessels with longer non-calcified segments and preserved distal coronary territory received higher scores, whereas anatomically unfavorable regions were penalized. The highest-scoring region was proposed as the optimal distal anastomosis target.

**Table 1:**
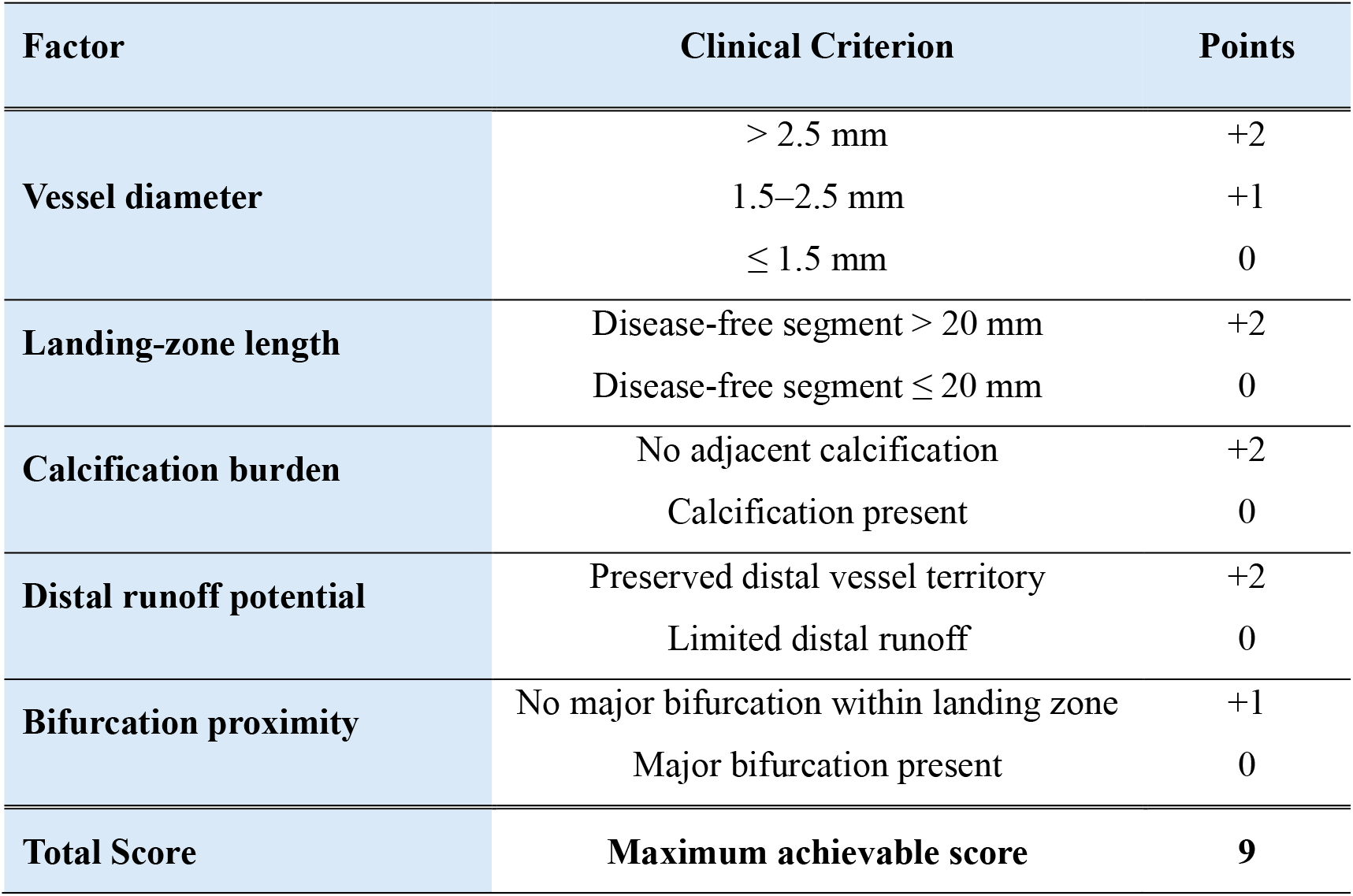
Clinical scoring framework for automated distal anastomosis site selection.

### 2.4. Interactive Bypass Graft Planning

Following identification of the optimal distal anastomosis target, a patient-specific bypass graft configuration is generated through an interactive planning framework integrating automated geometric modeling with surgeon-guided design control (Fig. 2). The surgeon retains full control over proximal and distal anastomosis location, conduit trajectory, and graft geometry, enabling rapid exploration of alternative revascularization strategies.

**Figure 2:**
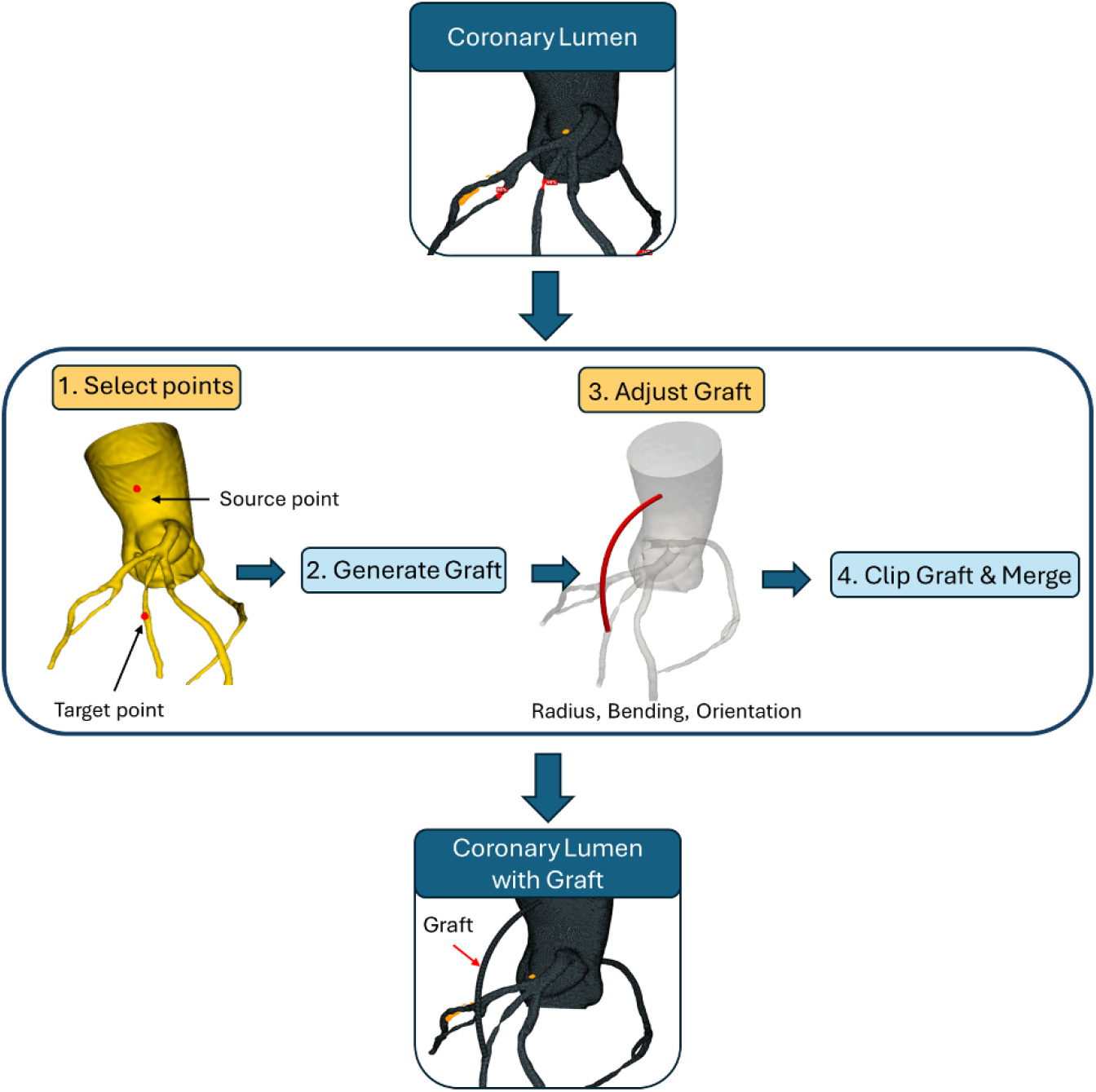
Overview of the interactive bypass graft planning framework. Automated components (blue) include centerline mapping, spline-based conduit generation, surface reconstruction, and coronary network integration. Interactive components (yellow) include surgeon-guided anastomosis selection and conduit geometry adjustment.

#### 2.4.1. Anastomosis Planning and Conduit Generation

The surgeon interactively selects proximal and distal anastomosis locations directly on the patient-specific coronary surface model. Selected points are automatically aligned to the nearest coronary centerline locations to maintain anatomical consistency with the native vascular geometry.

A smooth conduit trajectory is generated between the two anastomosis sites using a continuous spline representation. Graft shape is controlled through adjustable curvature and orientation parameters, allowing the conduit to be adapted to patient-specific cardiac anatomy and surrounding vascular structures. The resulting conduit is converted into a three-dimensional tubular graft surface with user-defined graft diameter, producing a closed and simulation-ready bypass geometry.

#### 2.4.2. Integration With the Coronary Network

Following graft generation, the bypass conduit is integrated with the native coronary arterial tree to produce a unified vascular network for hemodynamic simulation. The graft centerline is automatically extracted and connected to the corresponding coronary branches while preserving geometric continuity and vessel topology. Where required, short interpolation segments are introduced to ensure smooth transitions between the native coronary circulation and bypass conduit.

The final output consists of a continuous patient-specific coronary network incorporating both native coronary vessels and bypass grafts, which serves as the direct input for down-stream computational flow simulation and functional assessment, as detailed in Supplemental Material.

## 3. Results

### 3.1. Coronary Artery and Calcium Segmentation

The nnU-Net model was trained and evaluated using a five-fold cross-validation scheme on 20 coronary CTA scans annotated for both lumen and calcium. During validation, lumen segmentation was consistently excellent across all folds, yielding a mean DSC of 0.96 ± 0.03.

Calcium segmentation was more variable (mean DSC = 0.79 ± 0.03), reflecting the inherent challenge of detecting small, high-attenuation deposits embedded within heterogeneous vessel-wall tissue. The final model, retrained on all 20 training scans, was evaluated on eight previously unseen cases withheld from all training and tuning procedures. Lumen segmentation remained robust across all test cases (mean DSC = 0.96 ± 0.01). Calcium segmentation showed higher inter-case variability (mean DSC = 0.73 ± 0.15), with per-case scores ranging from 0.51 to 0.93.

### 3.2. Integrated Platform Performance and Time Efficiency

All five computational modules were integrated into a single interactive platform implemented in Python with a Qt-based graphical interface (PySide6) and PyVista/VTK-based 3D rendering. The interface organized workflow steps into a sequential button panel mirroring the clinical decision-making process, with all results automatically propagated between modules without manual file transfer.

End-to-end pipeline execution time from raw CCTA input to segmented anatomy, quantitative morphologic characterization, detected stenoses, optimal distal target selection, virtual graft geometry, and pre-/post-CABG FFR estimation was approximately seven minutes per case on a standard workstation. This represents a one to two order-of-magnitude reduction compared with conventional workflows requiring separate tools, manual segmentation, and computationally intensive 3D simulation. A detailed per-step timing comparison is summarized in Table 2.

**Table 2:** Analysis time comparison between conventional approaches and the integrated interactive platform. All automated steps were executed on a standard CPU workstation without high-performance computing resources.

| Coronary segmentation | Conventional CFD Workflow | Proposed Platform |
| --- | --- | --- |
| **Feature extraction** | 5–10 min | ~30 s |
| **Stenosis detection** | Visual inspection (subjective) | ~2 s |
| **CABG target identification** | Several minutes (manual review) | ~2 s |
| **Graft generation** | Not routinely available | ~2 min |
| **Hemodynamic analysis (FFR)** | Hours (3D CFD on HPC) | ~2 min |
| **Total pipeline** | **Several hours** | **~7 min** |

### 3.3. Comparison Between Simulated and Invasively Measured FFR Values

To assess the physiologic validity of the proposed platform, simulated FFR values were compared against invasively measured FFR in eight randomly selected patients from the study cohort. The validation subset included five lesions located in the LAD and three in the LCX, with measured FFR values spanning the clinically relevant range from 0.69 to 0.87 (Table 3).

**Table 3:** Comparison of platform-derived simulated FFR with reference measurements.

| Patient# | Stenosis Location | Simulated FFR | Measured FFR | Relative Difference (%) |
| --- | --- | --- | --- | --- |
| **P1** | LCX | 0.79 | 0.80 | 1.26 |
| **P2** | LAD | 0.81 | 0.82 | 1.20 |
| **P3** | LAD | 0.83 | 0.82 | 1.20 |
| **P4** | LAD | 0.79 | 0.79 | 0.00 |
| **P5** | LAD | 0.87 | 0.86 | 1.11 |
| **P6** | LCX | 0.82 | 0.85 | 3.58 |
| **P7** | LAD | 0.77 | 0.78 | 1.27 |
| **P8** | LCX | 0.69 | 0.72 | 4.25 |
| **Mean ± SD** |  |  |  | **1.73±1.42** |

Across all eight patients, the platform-derived FFR closely matched the reference measurements, with an overall mean absolute relative difference of 1.73 ± 1.42%. All individual relative differences were below 5%, demonstrating consistent agreement with invasive measurements across the validation subset.

The scatter plot of simulated against measured FFR (Figure 3) demonstrates a strong positive linear relationship between the two modalities, with data points clustering tightly around the line of identity. Pearson correlation analysis yielded a coefficient of r = 0.96 (R2 = 0.93), with a regression slope of 1.15 and intercept of −0.13.

**Figure 3:**
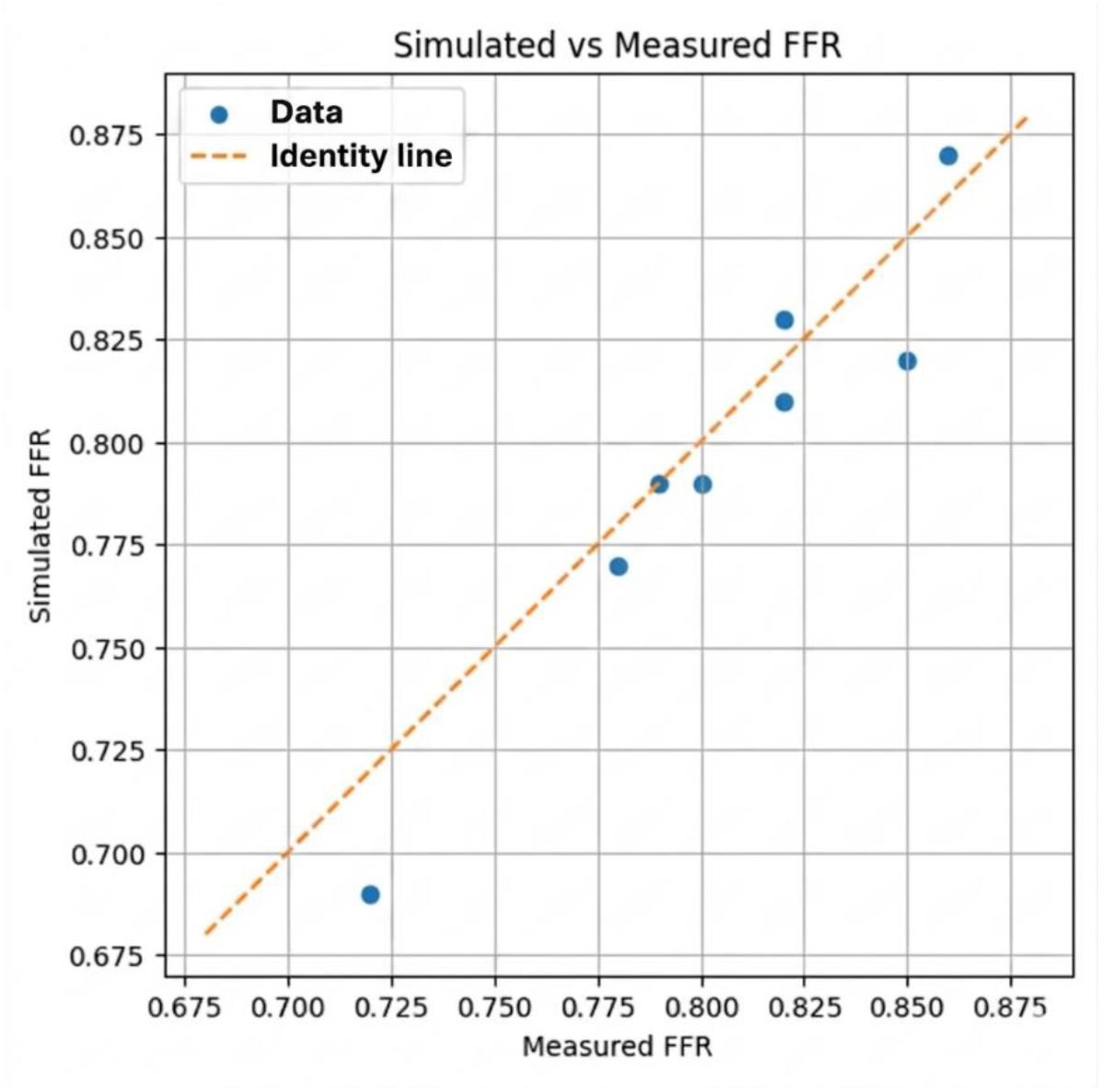
Validation of simulated FFR against invasive measurements. The dashed orange line denotes the line of identity (y = x).

## 4. Discussion

This study introduces a fully automated platform that integrates deep-learning segmentation, quantitative coronary morphology, automated stenosis detection, target-site scoring, interactive graft design, and rapid reduced-order hemodynamic modeling into a single pre-operative workflow for coronary artery bypass grafting. On held-out CCTA data, the pro-posed framework demonstrated the feasibility of integrating anatomical analysis, physiology-informed lesion assessment, and interactive bypass planning into a rapid and clinically tractable workflow. More importantly, the study highlights a broader conceptual shift in CABG planning from anatomy-driven interpretation toward quantitatively integrated surgical decision support. To our knowledge, this is the first study to combine automated coronary segmentation, quantitative target-site analysis, virtual graft generation, and reduced-order hemodynamic simulation within a unified CABG planning environment.

### 4.1. Context in literature

Our lumen Dice coefficient is at the upper end of recently reported coronary segmentation performance [8, 12], reflecting the robustness of the nnU-Net self-configuring framework. Calcium segmentation performance remained within the range reported for this substantially more challenging task, where blooming artifact, partial-volume effects, and heterogeneous contrast continue to limit segmentation accuracy [13, 14, 15]. More broadly, this work reflects an ongoing transition in cardiovascular imaging from descriptive visualization toward computationally augmented procedural planning. Although coronary CTA has become increasingly established as a noninvasive modality for anatomic coronary assessment, translation of these datasets into actionable surgical guidance has remained limited by fragmented workflows, computational burden, and dependence on manual interpretation. Existing CT-derived FFR frameworks primarily focus on lesion-level physiologic classification, whereas surgical planning requires a substantially broader understanding of coronary anatomy, distal vessel quality, graft geometry, runoff potential, and competing flow physiology. Moreover, classical 3D computation-based CT-FFR requires off-site high-performance computing with reported times of 1 to 4 hours per case [16, 17], and patient-specific bypass simulation remains impractical for routine preoperative use [9]. Reduced-order and lumped-parameter hemodynamic formulations have helped address some of these computational limitations by recovering clinically relevant pressure and flow characteristics at a fraction of the cost of full 3D simulations, enabling near-real-time hemodynamic assessment [18, 19, 20]. However, most prior work has remained isolated to either CT-FFR estimation or standalone computational bypass simulation without integration into a cohesive surgical workflow.

A central contribution of the present study is therefore not solely the performance of the individual computational modules, but the integration of these disconnected components into a unified and interactive planning framework. The FASTTRACK CABG study has explored the feasibility of performing surgical planning using CCTA and CT-derived FFR without invasive angiography [7]; however, that approach still depends on manual segmentation, semi-manual morphometry, and separate hemodynamic analysis. The present framework extends this concept toward a more automated and quantitatively standardized planning environment that may reduce operator variability, improve reproducibility of target selection, and facilitate broader adoption of physiology-informed surgical planning beyond highly specialized computational centers.

### 4.2. Clinical implications

For the operating surgeon, several aspects of the proposed framework are of immediate practical relevance. First, stenosis severity, landing-zone quality, and distal perfusion are quantified using objective descriptors, addressing the known inter-observer variability associated with visual angiographic assessment [4, 21]. Importantly, the proposed framework attempts to formalize aspects of CABG planning that are traditionally experienced-driven and only qualitatively assessed. In routine practice, surgeons mentally integrate vessel caliber, plaque burden, distal runoff, conduit trajectory, and anticipated competitive flow when selecting bypass targets. Much of this reasoning is implicit, variable between operators, and difficult to standardize across institutions or training levels. By translating these considerations into quantitative descriptors and transparent scoring criteria, the platform provides a reproducible framework that may support surgical consistency while preserving physician oversight and flexibility. Third, the interactive graft-planning environment also enables iterative evaluation of alternative revascularization strategies in a manner that is difficult to achieve using conventional static imaging review alone. This capability may be particularly valuable in anatomically complex multivessel disease, diffuse distal atherosclerosis, redo operations, or sequential grafting strategies, where target selection and conduit configuration are frequently nuanced and highly operator-dependent.

From a workflow perspective, reducing total computation time from several hours to approximately seven minutes may represent a key requirement for clinical adoption. A planning framework that can be executed during a routine preoperative case-review meeting is substantially more compatible with real-world surgical workflow than approaches requiring dedicated simulation expertise and prolonged offline computation.

### 4.3. Limitations

Several limitations should be acknowledged. The segmentation network was trained on 20 annotated CCTA studies and tested on 8 held-out cases from a single institution; therefore, generalizability across scanners, reconstruction protocols, and patient populations remains to be established through larger multicenter validation studies. Calcium segmentation, although sufficient for downstream analysis, demonstrated greater variability, with smallest or more diffuse calcified deposits being the most likely to be under detected. Most importantly, neither the distal anastomosis recommendations nor the post-bypass FFR predictions have yet been validated against surgeon consensus, postoperative graft patency, invasive post-bypass physiologic assessment, or long-term clinical outcomes such as major adverse cardiac events. The platform should therefore be interpreted at present as a decision-support tool rather than definitive clinical guidance. Future work should therefore focus not only on technical validation, but also on demonstrating clinical utility, including agreement with expert surgical planning, impact on operative decision making, graft patency, procedural reproducibility, and downstream patient outcomes.

Accordingly, the platform is intentionally designed as a surgeon-in-the-loop decision-support system rather than an autonomous planning tool, preserving clinical judgment while augmenting quantitative decision support.

## 5. Conclusion

The proposed workflow integrates automated coronary segmentation, quantitative analytical and physiologic analysis, interactive virtual graft planning, and rapid reduced-order hemodynamic simulation into a unified platform that converts routine CCTA into a patient-specific CABG planning strategy in approximately seven minutes. By combining anatomical characterization, quantitative target-site analysis, virtual graft planning, and physiologic assessment within a clinically practical timeframe, the framework advances CABG planning toward a more standardized, reproducible, and physiology-informed paradigm. Although prospective multicenter validation and outcome-based evaluation remain necessary, the proposed system establishes a foundation for integrating AI-driven computational planning into routine surgical decision making and next-generation image-guided coronary revascularization.

## Data Availability

All data produced in the present study are available upon reasonable request to the authors

## Acknowledgment

FRN gratefully acknowledges Career Award from American Heart Association (25CDA1452999). The authors used ChatGPT (OpenAI) to assist with grammar and readability of the manuscript; all content was subsequently reviewed and edited by the authors, who take full responsibility for the work.

## Data Availability

Imaging data were acquired at Brigham and Women’s hospital (Boston, USA). Derived data supporting this study could be made available by the corresponding author (MR) upon reasonable request and in accordance with institutional policy.

