## Supplemental Material for "From CCTA to Surgical Strategy: An Integrated AI Framework for Patient-Specific Coronary artery bypass grafting Planning"

### Segmentation Model Training and Evaluation

Segmentation of the coronary tree and calcium deposits was carried out with the nnU-Net framework[1], which tailors preprocessing, patch size, voxel spacing, intensity normalization, and data augmentation to the characteristics of the input dataset. The network was trained on the manually annotated lumen and calcium masks. Training proceeded for up to 500 epochs, each corresponding to a full pass over the training patches. Throughout training, early stopping triggered when the validation loss and class-specific Dice scores no longer improved.

The 15 training/validation scans were used in a five-fold cross-validation setup to assess model stability. Following cross-validation, a final model was retrained on the full training/validation cohort and evaluated on to the 5 held-out test cases.

Segmentation performance was measured with the Dice Similarity Coefficient (DSC) [2], which for a ground-truth voxel set $A$ and a predicted voxel set $B$ is given by:

$$\mathrm{DSC}(A, B) = \frac{2\,|A \cap B|}{|A| + |B|},$$

with a DSC of 1 indicating perfect overlap between the two masks. In addition to voxel-wise overlap metrices, all segmentations underwent visual review to assess topological integrity and identify anatomically implausible predictions not captured by DSC alone.

### Supplementary Feature Definitions

Table 1: Global quantification features of coronary calcium.

| **Feature** | **Definition** |
|---|---|
| Number of Deposits [count] | Total number of disconnected calcium components (connected regions in the mask). |
| Total Volume [$mm^3$] | Sum of volumes of all deposits. |
| Total Surface Area [$mm^2$] | Estimated surface area of all deposits from mesh triangulation. |
| Number of Calcified Segments [count] | Number of coronary artery branches containing at least one deposit. |

*Supplementary Table S2: Calcium Shape Features*

Table 2: Morphological features of calcium deposits.

| **Feature** | **Definition / Formula** |
|---|---|
| Volume [$mm^3$] | Voxel count $\times$ voxel volume. |
| Surface Area [$mm^2$] | Surface from marching-cubes triangulation. |
| Surface Area-to-Volume Ratio (SVR) [–] | $$\mathrm{SVR} = \frac{A}{V}$$ with $A$ = surface, $V$ = volume. |
| Sphericity [–] [3] | $$\Psi = \frac{\pi^{1/3}(6V)^{2/3}}{A}, \quad \Psi = 1 \text{ for a sphere.}$$ |
| Elongation [–] [3] | $$\mathrm{Elong} = \frac{\lambda_2}{\lambda_1}$$ ratio of principal inertia eigenvalues ($\lambda_1 \geq \lambda_2 \geq \lambda_3$). |
| Flatness [–] [3] | $$\mathrm{Flat} = \frac{\lambda_3}{\lambda_1}$$ ($\lambda_i$ = eigenvalues of inertia tensor, reflecting spread of voxel coordinates). |
| Mean HU [HU] | Mean CT intensity inside deposit. |

*Supplementary Table S3: Lumen Shape Features*

Table S3. Global lumen features (length-weighted means across branches).

| **Feature** | **Definition / Formula** |
|---|---|
| NumSegments [count] | Number of extracted branches. |
| NumBifurcations [count] | Nodes with degree > 2 in centerline graph. |
| TotalLength [mm] | $$L_{\mathrm{tot}} = \sum_b L_b$$ $L_b$ = length of branch $b$. |
| MeanTortuosity [–] | $$\mathrm{Tort}_b = \frac{L_b}{d_b}, \quad \overline{\mathrm{Tort}} = \frac{\sum_b L_b \cdot \mathrm{Tort}_b}{\sum_b L_b}$$ $d_b$ = Euclidean end-to-end distance of branch $b$. |
| MeanCurvature [$\mathrm{mm}^{-1}$] [4] | $$\kappa = \frac{\lVert \mathbf{r}' \times \mathbf{r}'' \rVert}{\lVert \mathbf{r}' \rVert^3}$$ |
| MeanTorsion [$\mathrm{mm}^{-1}$] [4] | $$\tau = \frac{(\mathbf{r}' \times \mathbf{r}'') \cdot \mathbf{r}'''}{\lVert \mathbf{r}' \times \mathbf{r}'' \rVert^2}$$ $\mathbf{r}(s)$: centerline position parameterized by arclength $s$; primes denote derivatives. |
| MeanRadius [mm] | Length-weighted average of local lumen radii. |
| MeanBranchAngle [°] | $$\theta = \cos^{-1}\left(\frac{v_{\mathrm{parent}} \cdot v_{\mathrm{child}}}{\lVert v_{\mathrm{parent}} \rVert \lVert v_{\mathrm{child}} \rVert}\right)$$ |
| MeanBifAngle [°] | Mean daughter–daughter angle at all bifurcation nodes. |

*Supplementary Table S4: Calcium Topology Features*

*Supplementary Table S5: Lumen–Calcium Interaction Features*

Table S4. Topological features of coronary calcium.

| **Feature** | **Definition** |
|---|---|
| Sparsity [$\text{mm}^{-3}$] | Mean pairwise centroid distance normalized by mean deposit volume. |
| Min Distance to Bifurcation [mm] | Euclidean distance from deposit centroid to nearest bifurcation node. |
| Dominant Segment [categorical] | Branch (LMCA, LAD, LCx, RCA) containing the largest fraction of calcium by volume. |
| Distribution per Segment [%] | Relative voxel percentage of calcium per coronary segment. |
| Anatomical Side [categorical] | "Myocardial" or "Pericardial" based on deposit centroid orientation relative to vessel centerline. |

Table S5. Interaction features between lumen and calcium.

| **Feature** | **Definition / Formula** |
|---|---|
| Min Distance to Lumen [mm] | $$d_{\min} = \min_{p \in \partial C,\, q \in \partial L} \lVert p - q \rVert$$ $\partial C$, $\partial L$: calcium and lumen surfaces, respectively. |
| Contact Ratio [–] | $$\text{CR} = \frac{N_{\text{contact}}}{N_{\text{surface}}}$$ $N_{\text{contact}}$: calcium surface points within 0.5 mm of the lumen boundary. |
| Arc Angle [°] | Angular span of the deposit projected onto the cross-sectional plane normal to the local vessel tangent. |

### Hemodynamic Assessment Along the Coronary Centerline via Distributed Lumped Parameter Modeling

The present framework adopts a Distributed Lumped Parameter (DLP) model to compute coronary hemodynamics along the centerline network. Consistent with established one-dimensional hemodynamic formulations [5], the governing equations are expressed in terms of the cross-sectionally averaged pressure $P(x,t)$ and volumetric flow rate $Q(x,t)$, where $x$ is the axial coordinate along a vessel segment and $t$ denotes time. Conservation of mass is enforced at each bifurcation node as $\sum_i Q_i^{\text{in}} = \sum_j Q_j^{\text{out}}$, while the momentum balance along a segment of length $L$ takes the integrated form

$$\frac{\rho L}{\pi \int_0^1 R(x)^2 \, dx} \frac{\partial Q}{\partial t} + \mathcal{R} Q + \Delta P = 0, \tag{1}$$

where $\rho$ is the blood density, $R(x)$ is the lumen radius, and $\Delta P = P_{\text{out}} - P_{\text{in}}$ is the pressure drop across the segment. The inertial term is captured by the first expression, and $\mathcal{R}$ is a generalized flow resistance decomposed as

$$\mathcal{R} = \mathcal{R}_v + \mathcal{R}_c + \mathcal{R}_e,$$

corresponding to viscous (Poiseuille) losses, convective acceleration losses arising from axial radius variation, and expansion losses distal to abrupt geometric narrowings, respectively.

Hemodynamic quantities, specifically pressure, flow rate, and fractional flow reserve (FFR), were evaluated at end-diastole, a phase during which coronary flow is approximately steady. Under the quasi-steady assumption, the temporal derivative was neglected ($\partial Q/\partial t = 0$), reducing Eq. (1) to a purely resistive pressure–flow relationship:

$$\mathcal{R} Q + \Delta P = 0. \tag{2}$$

Blood was modeled as a Newtonian fluid with density $\rho = 1060\,\text{kg/m}^3$ and dynamic viscosity $\mu = 0.0035\,\text{Pa·s}$, consistent with standard assumptions for large-vessel coronary flow analysis. A detailed derivation of the resistance terms and the network assembly procedure is provided in Supplementary Material S1.

Applied to each segment of the extracted centerline tree and coupled through junction continuity conditions, Eq. (2) yields a linear system in the nodal pressures and segmental flow rates. Patient-specific aortic inflow was prescribed at the inlet, while distal microvascular boundary conditions were represented by lumped terminal resistances scaled to the myocardial territory subtended by each outlet branch and calibrated to reproduce hyperemic flow conditions consistent with clinical FFR measurements. For truncated vascular tree, outlet resistances were assigned based on Murray's law scaling to preserve physiologic flow distribution. Solving the assembled linear system yielded nodal pressure

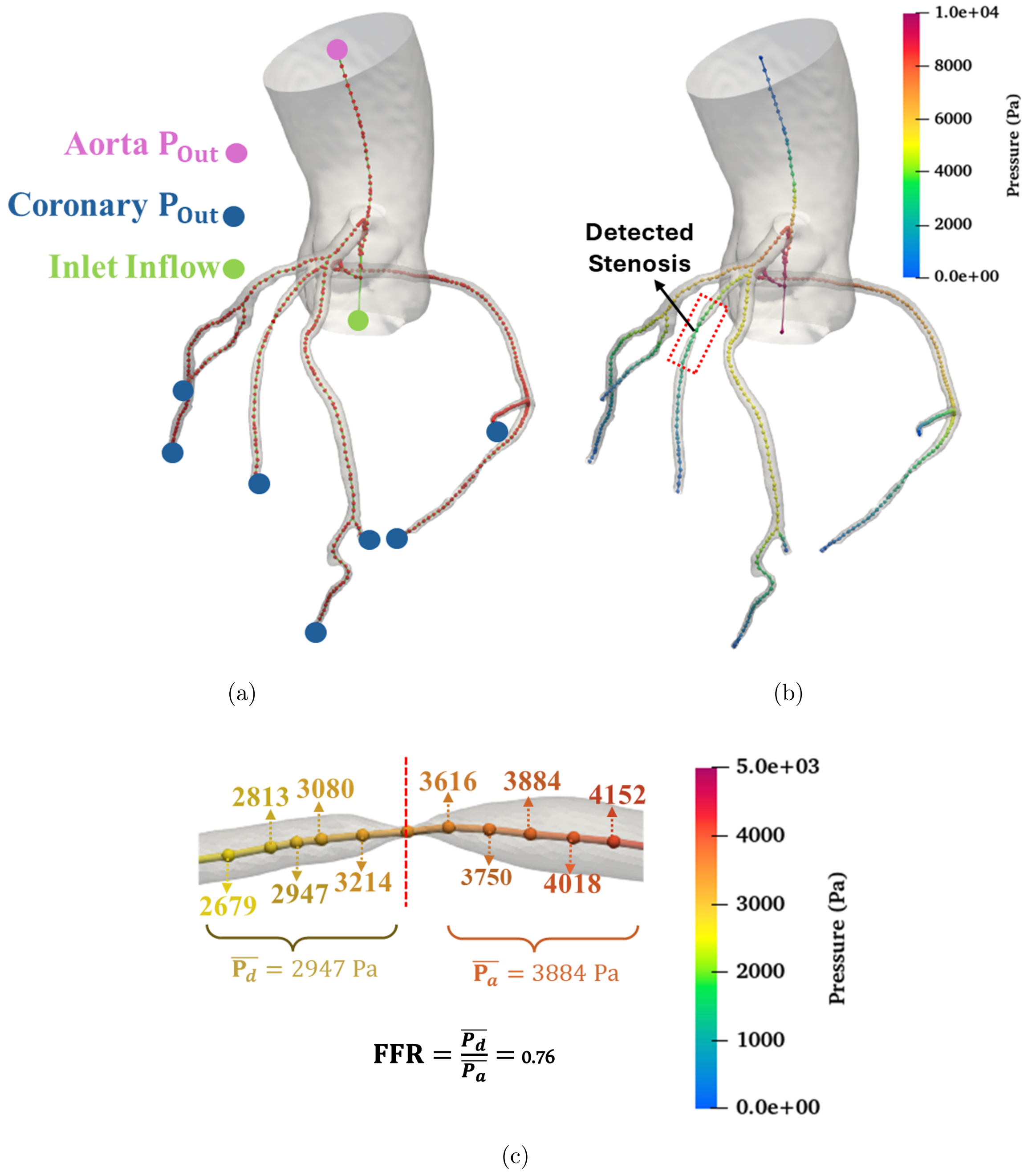


Figure 1: Illustration of the DLP-based hemodynamic assessment pipeline. (a) Extracted coronary centerline tree with annotated stenotic segments. (b) Pressure distribution computed via the DLP model under hyperemic boundary conditions. (c) FFR values derived at each stenosis, demonstrating the predicted hemodynamic impact of the planned bypass graft.

and segmental flow rate throughout the centerline tree.

As illustrated in Figure 1, the computed centerline pressure field is subsequently used to derive FFR at each identified stenosis. Following stenosis localization (Section **??**), the mean proximal aortic pressure $\bar{P}_a$ and the mean distal coronary pressure $\bar{P}_d$ were computed by averaging the DLP-derived pressure over the five centerline nodes immediately proximal and distal to the stenosis, respectively, to suppress local numerical noise. FFR was then computed according to its standard hemodynamic definition:

$$\mathrm{FFR} = \frac{\bar{P}_d}{\bar{P}_a}.$$
